# Effects of temperature variation and humidity on the mortality of COVID-19 in Wuhan

**DOI:** 10.1101/2020.03.15.20036426

**Authors:** Yueling Ma, Yadong Zhao, Jiangtao Liu, Xiaotao He, Bo Wang, Shihua Fu, Jun Yan, Jingping Niu, Bin Luo

## Abstract

**Object:** Meteorological parameters are the important factors influencing the infectious diseases like severe acute respiratory syndrome (SARS). This study aims to explore the association between coronavirus disease (COVID-19) death and weather parameters.

**Methods:** In this study, we collected the daily death number of COVID-19, meteorological and air pollutant data from 20 January, 2020 to 29 February, 2020 in Wuhan, China. Then, the generalized additive model was applied to explore the impact of temperature, humidity and diurnal temperature range on daily mortality of COVID-19.

**Results:** There were in total 2299 COVID-19 mortality counts in Wuhan. A positive association with COVID-19 mortality was observed for diurnal temperature range (r = 0.44), but negative association for relative humidity (r = −0.32). In addition, each 1 unit increase in diurnal temperature range was only associated with a 2.92% (95% CI: 0.61%, 5.28%) increase in COVID-19 mortality at lag 3. However, both per 1 unit increase of temperature and absolute humidity were related to the decreased COVID-19 mortality at lag 3 and lag 5, respectively.

**Conclusion:** In summary, this study suggests the temperature variation and humidity may be important factors affecting the COVID-19 mortality.

## 1. Introduction

In December 2019, a novel coronavirus disease (COVID-19) broke out in Wuhan, China, which caused by severe acute respiratory syndrome coronavirus 2 (SARS-CoV-2) [1, 2]. The COVID-19 had been affirmed to have human-to-human transmissibility [3] and has now spread worldwide, which raised high attention not only within China but internationally. The World Health Organization (WHO) reported that there are 118, 326 confirmed cases and 4292 deaths in globally until March 11, 2020 [4], and evaluated as global pandemic on the same day [5].

In retrospect studies, the outbreak of severe acute respiratory syndrome (SARS) in Guangdong in 2003 gradually faded with the warming weather coming, and was basically ended until July [6]. It has been documented that the temperature and its variations might have affected the SARS outbreak [7]. A study in Korea found that the risk of influenza incidence was significantly increased with low daily temperature and low/high relative humidity, a positive significant association was observed for diurnal temperature range (DTR) [8]. Moreover, temperature [9] and DTR [10] have been linked to the death from respiratory diseases. A study demonstrated that absolute humidity had significant correlations with influenza viral survival and transmission rates [11]. Couple of studies reported that the COVID-19 was related to the meteorological factors, which decreased with the temperature increasing [12, 13], but their effects on the mortality had not been reported. Therefore, we assume that the weather conditions might also contributed to the mortality of COVID-19.

As the capital of Hubei Province and one of the largest city in central China, Wuhan is located in the middle of the Yangtze River Delta, which has a typical subtropical, humid, monsoon climate with cold winters and warm summers [14]. The average annual temperature and rainfall are 15.8 °C–17.5 °C and 1050 mm–2000 mm, respectively [15]. Besides, Wuhan owns an area of 8494 km^2^ and a population over 11 million (as of 2018) [16]. So far, this COVID-19 has caused 5393 deaths globally, and caused 2456 deaths in Wuhan. Although the deaths may be affected by many factors, the present is to explore the effect from meteorological angel with generalized additive model (GAM).

## 2. Methods

### 2.1. Data collection

Data from 20 January to 29 February 2020 in Wuhan were collected, including daily death number of coronavirus disease (COVID-19), meteorological, and air pollutant data. Daily death number of COVID-19 were collected from the official website of Health Commission of Hubei Province, people’s Republic of China (http://wjw.hubei.gov.cn/). The simultaneous daily meteorological and air pollutant data were obtained from Shanghai Meteorological Bureau and Data Center of Ministry of Ecology and Environment of the People’s Republic of China, respectively. Meteorological variables included daily average temperature, diurnal temperature range (DTR) and relative humidity, and air pollutant data included particulate matter with aerodynamic diameter ≤10 μm (PM_10_), particulate matter with aerodynamic diameter ≤2.5 μm (PM_2.5_), nitrogen dioxide (NO_2_), and sulfur dioxide (SO_2_), carbon monoxide (CO), ozone (O_3_). Therefore, there is no need to have ethical review.

### 2.2. Calculation of absolute humidity

Absolute humidity was calculated according to the previous study and was measured by vapor pressure (VP) [17]. The density of water vapor, or absolute humidity [ρ_*v*_ (g/m^3^)], is the mass of moisture per total volume of air. It is associated to VP via the ideal gas law for the moist portion of the air:

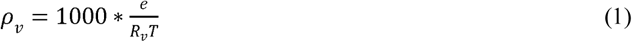

Where *e* is vapor pressure (VP), R_*v*_ is the gas constant for water vapor [461.53 J/(kg K)], and T is the daily ambient temperature (K). VP is a commonly used indicator of absolute humidity and is calculated from ambient temperature and relative humidity using the Clausius–Clapeyron relation [18]. Briefly, we first calculated the saturation vapor pressure [e_s_ (T) (mb)] from daily ambient temperature using the following equation:

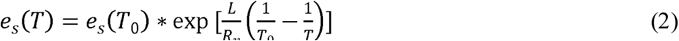

Where e_*s*_ (T_0_) denotes saturation vapor pressure at a reference temperature T_0_ (273.15 K) which equals to 6.11 mb. L denotes the latent heat of evaporation for water (2257 kJ/kg). R_*v*_ is the gas constant for water vapor [461.53 J/(kg K)]. T denotes daily ambient temperature (K). Then, VP (Pa) was calculated by combining the e_*s*_ (T) calculated using Eq. (2) with relative humidity (RH):

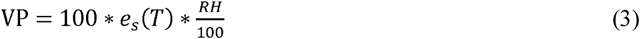

### 2.3. Statistical methods

A descriptive analysis was performed. We used a generalized additive model (GAM) to analyze the associations between meteorological factors (temperature, DTR, relative humidity and absolute humidity) and the daily mortality of COVID-19. The core analysis was a GAM with a quasi-Poisson link function based on the previous studies [19, 20]. We first built the basic models for mortality outcomes without including air pollution or weather variables. We incorporated smoothed spline functions of time, which accommodate nonlinear and nonmonotonic patterns between mortality and time, thus offering a flexible modeling tool. Then, we introduced the weather variables and analyzed their effects on mortality. Akaike’s information criterion was used as a measure of how well the model fitted the data. Consistent with several recent time-series studies [21, 22], the penalized smoothing spline function was applied to control the effects of confounding factors, such as time trends, day-of-week and air pollution. The core GAM equation is:

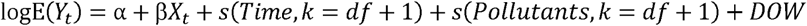

Where t is the day of the observation; E(Y_t_) is the expected number of daily mortality for COVID-19 on day t; *α* is the intercept; β is the regression coefficient; X_t_ is the daily level of weather variables on day t; s() denotes the smoother based on the penalized smoothing spline. Based on quasi Akaike information criterion (QAIC), the 2 degrees of freedom (df) was used for time trends and 3 df for air pollutants, temperature and relative humidity; DOW is a categorical variable indicating the date of the week.

After establishing the core model, we also considered the lag effects of weather conditions on death of COVID-19, and examined the potentially lagged effects, i.e., single day lag (from lag 0 to lag 5) and multiple-day average lag (from lag 01 to lag 05) [23]. The exposure and response correlation curves between weather variables and COVID-19 mortality were plotted using a spline function in the GAM. We also performed a sensitivity analysis by changing the df of the penalized smoothing spline function from 2 to 9 for calendar time and from 3 to 8 for temperature and humidity.

All of the statistical analyses were two-sided, and at a 5% level of significance. All analyses were conducted using R software (version 3.5.3) with the GAM fitted by the “mgcv” package (version 1.8-27). The effect estimates were expressed as the percentage changes and their 95% confidence intervals (CIs) in daily mortality of COVID-19 associated with per 1 unit increase in weather variables.

## 3. Results

### 3.1. Descriptive results of COVID-19 mortality, meteorological variables and air pollutants

Table 1 showed the descriptive statistics for daily deaths of COVID-19, meteorological variables and air pollutants. Over the observation period (January 20, 2020 to February 29, 2020), there were 2,299 COVID-19 deaths in Wuhan. On average, there were approximately 56 deaths of COVID-19 per day. Temperatures ranged from 1.8 °C to 18.7 °C, and DTR ranged from 2 °C to 17.5 °C. Average temperature and DTR during this period were 7.44°C and 9.15 °C, respectively. The relative humidity and absolute humidity were 59%-97% with an average 82.24% and 4.27 g/m^3^-11.63 g/m^3^ with an average 6.69 g/m^3^, respectively. The mean concentrations of PM_2.5_, PM_10_, NO_2_, SO_2_, O_3_, and CO were 44.68 μg/m^3^, 52.56 μg/m^3^, 23.02 μg/m^3^, 7.29 μg/m^3^, 73.76 μg/m^3^ and 0.91 mg/m^3^, respectively.

**Table 1.** Summary of COVID-19 mortality counts, meteorological data and air pollutants

| Variables | Daily measures |  |  |  |  |  |
| --- | --- | --- | --- | --- | --- | --- |
| | Mean ± S.D. | Min | P<sub>25</sub> | Median | P<sub>75</sub> | Max |
| Mortality counts | 56.07 ± 42.69 | 2.00 | 25.00 | 49.00 | 76.00 | 216.00 |
| Meteorological factors |  |  |  |  |  |  |
| Temperature (°C) | 7.44 ± 3.96 | 1.80 | 4.40 | 6.50 | 9.90 | 18.70 |
| DTR (°C) | 9.15 ± 4.74 | 2.00 | 4.70 | 8.70 | 14.00 | 17.50 |
| Relative humidity (%) | 82.24 ± 8.51 | 59.00 | 77.00 | 83.00 | 88.00 | 97.00 |
| Absolute humidity (g/m<sup>3</sup>) | 6.69 ± 1.78 | 4.27 | 5.38 | 6.30 | 7.51 | 11.63 |
| Concentrations of air pollutants |  |  |  |  |  |  |
| PM<sub>2.5</sub> (µg/m<sup>3</sup>) | 44.68 ± 23.97 | 9.00 | 28.00 | 41.00 | 64.00 | 97.00 |
| PM<sub>10</sub> (µg/m<sup>3</sup>) | 52.56 ± 26.01 | 12.00 | 32.00 | 48.00 | 69.00 | 116.00 |
| NO<sub>2</sub> (µg/m<sup>3</sup>) | 23.02 ± 11.77 | 10.00 | 16.00 | 20.00 | 28.00 | 76.00 |
| SO<sub>2</sub> (µg/m<sup>3</sup>) | 7.29 ± 2.28 | 5.00 | 5.00 | 7.00 | 9.00 | 13.00 |
| CO (mg/m<sup>3</sup>) | 0.91 ± 0.21 | 0.50 | 0.80 | 0.90 | 1.00 | 1.40 |
| O<sub>3</sub> (µg/m<sup>3</sup>) | 73.76 ± 21.50 | 39.00 | 53.00 | 74.00 | 94.00 | 110.00 |

COVID-19, coronavirus disease; SD, standard deviance; Min, Minimum; P<sub>25</sub>, 25th percentile; P<sub>75</sub>, 75th percentile; Max, Maximum; DTR, diurnal temperature range; PM<sub>2.5</sub>, particulate matter with aerodynamic diameter $\leq 2.5$ µm; PM<sub>10</sub>, particulate matter with aerodynamic diameter $\leq 10$ µm; NO<sub>2</sub>, nitrogen dioxide; SO<sub>2</sub>, sulfur dioxide; CO, carbon monoxide; O<sub>3</sub>, ozone.

Fig. 1 presented the temporal pattern of daily mortality counts of COVID-19 and meteorological factor levels in the study period, showing the daily death number of COVID-19 had a similar pattern with temperature and absolute humidity.

**Fig. 1.**
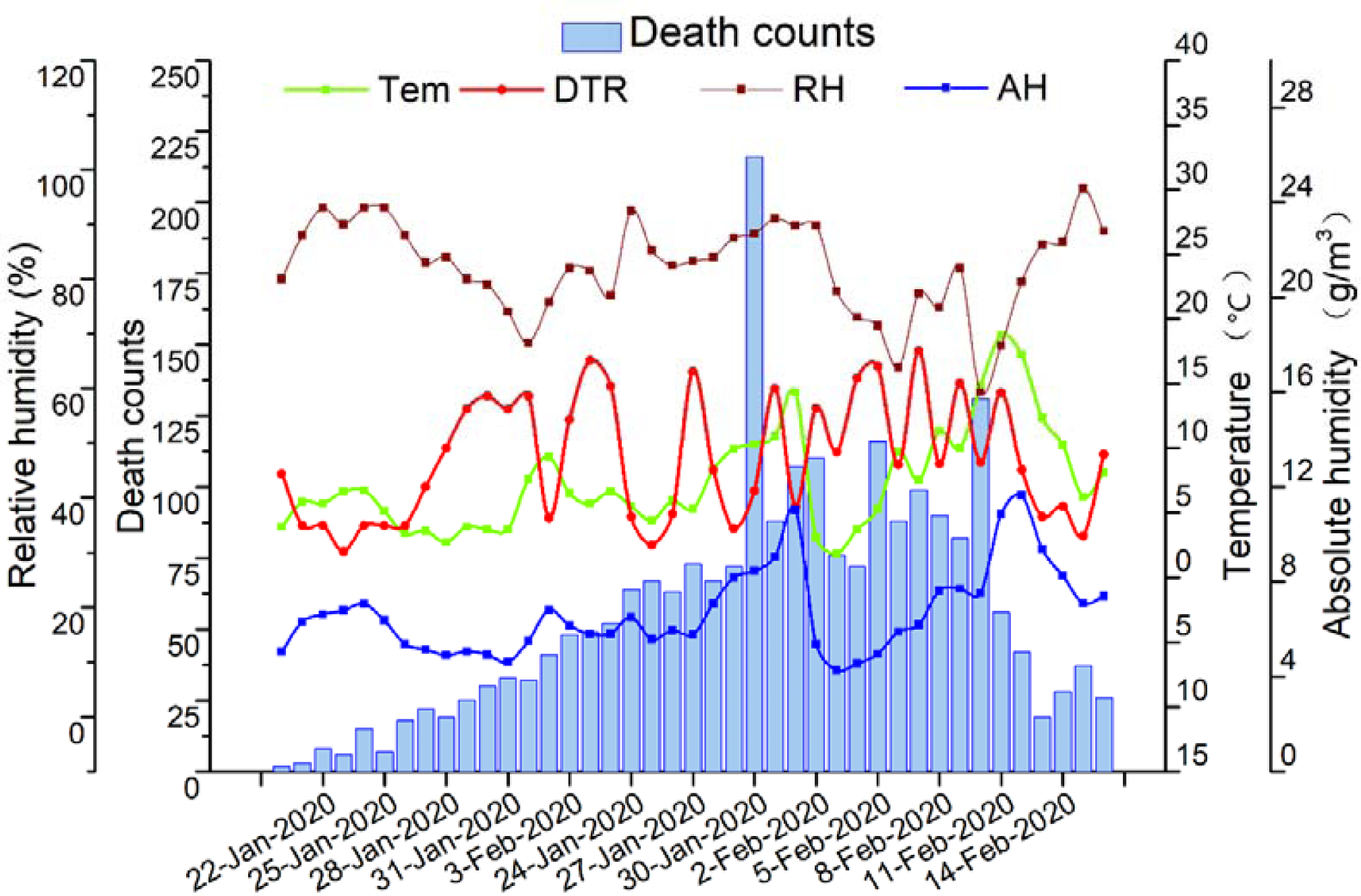
Temporal pattern of COVID-19 daily mortality and meteorological factor levels in Wuhan, China, from 20 January to 29 February 2020. Tem: temperature; DTR, diurnal temperature range; RH: relative humidity; AH: absolute humidity.

### 3.2. Correlation between COVID-19 mortality and meteorological factors and air pollutants

The correlation results between mortality counts of COVID-19, meteorological measures and air pollutant concentrations were presented in Table 2. The mortality counts of COVID-19 was negatively associated with relative humidity (r = −0.32), PM_2.5_ (r = −0.53) and PM_10_ (r = −0.45). A positive association with COVID-19 mortality was observed for DTR (r = 0.44), and SO_2_ (r = 0.31).

**Table 2.** Spearman’s correlation between meteorological factors and air pollutants and COVID-19 mortality

|  | Mortality | Tem | DTR | RH | AH | PM<sub>2.5</sub> | PM<sub>10</sub> | NO<sub>2</sub> | SO<sub>2</sub> | CO | O<sub>3</sub> |
| --- | --- | --- | --- | --- | --- | --- | --- | --- | --- | --- | --- |
| Mortality | 1.00 |  |  |  |  |  |  |  |  |  |  |
| Tem | 0.30 | 1.00 |  |  |  |  |  |  |  |  |  |
| DTR | 0.44* | -0.06 | 1.00 |  |  |  |  |  |  |  |  |
| RH | -0.32* | -0.08 | -0.59* | 1.00 |  |  |  |  |  |  |  |
| AH | 0.16 | 0.90* | -0.30 | 0.31* | 1.00 |  |  |  |  |  |  |
| PM<sub>2.5</sub> | -0.53* | 0.03 | 0.02 | -0.20 | -0.05 | 1.00 |  |  |  |  |  |
| PM<sub>10</sub> | -0.45* | 0.11 | 0.06 | -0.25 | 0.01 | 0.97* | 1.00 |  |  |  |  |
| NO<sub>2</sub> | -0.04 | 0.21 | 0.33* | -0.39* | -0.03 | 0.63* | 0.65* | 1.00 |  |  |  |
| SO<sub>2</sub> | 0.31* | 0.41* | 0.59* | -0.69* | 0.08 | 0.31* | 0.40* | 0.71* | 1.00 |  |  |
| CO | -0.06 | 0.57* | 0.08 | -0.01 | 0.51* | 0.52* | 0.56* | 0.59* | 0.52* | 1.00 |  |
| O<sub>3</sub> | 0.21 | 0.03 | 0.75* | -0.80* | -0.26 | 0.22 | 0.29 | 0.33* | 0.62* | 0.01 | 1.00 |

COVID-19, coronavirus disease; Tem, temperature; DTR, diurnal temperature range; RH, relative humidity; AH, absolute humidity; PM<sub>2.5</sub>, particulate matter with aerodynamic diameter $\leq 2.5$ µm; PM<sub>10</sub>, particulate matter with aerodynamic diameter $\leq 10$ µm; NO<sub>2</sub>, nitrogen dioxide; SO<sub>2</sub>, sulfur dioxide; CO, carbon monoxide; O<sub>3</sub>, ozone. \* $P < 0.05$ .

### 3.3. Effects of temperature, humidity and DTR on COVID-19 mortality

Fig. 2 flexibly plotted the exposure-response relationship curves between meteorological factors and COVID-19 mortality at current day. Generally, the curves tended to be not associated with COVID-19 mortality for DTR but were strongly positive for temperature. In addition, the curves associated with relative humidity and absolute humidity presented similar linear trends, which indicated that the higher level of humidity might cause decrease in the COVID-19 mortality.

**Fig. 2.**
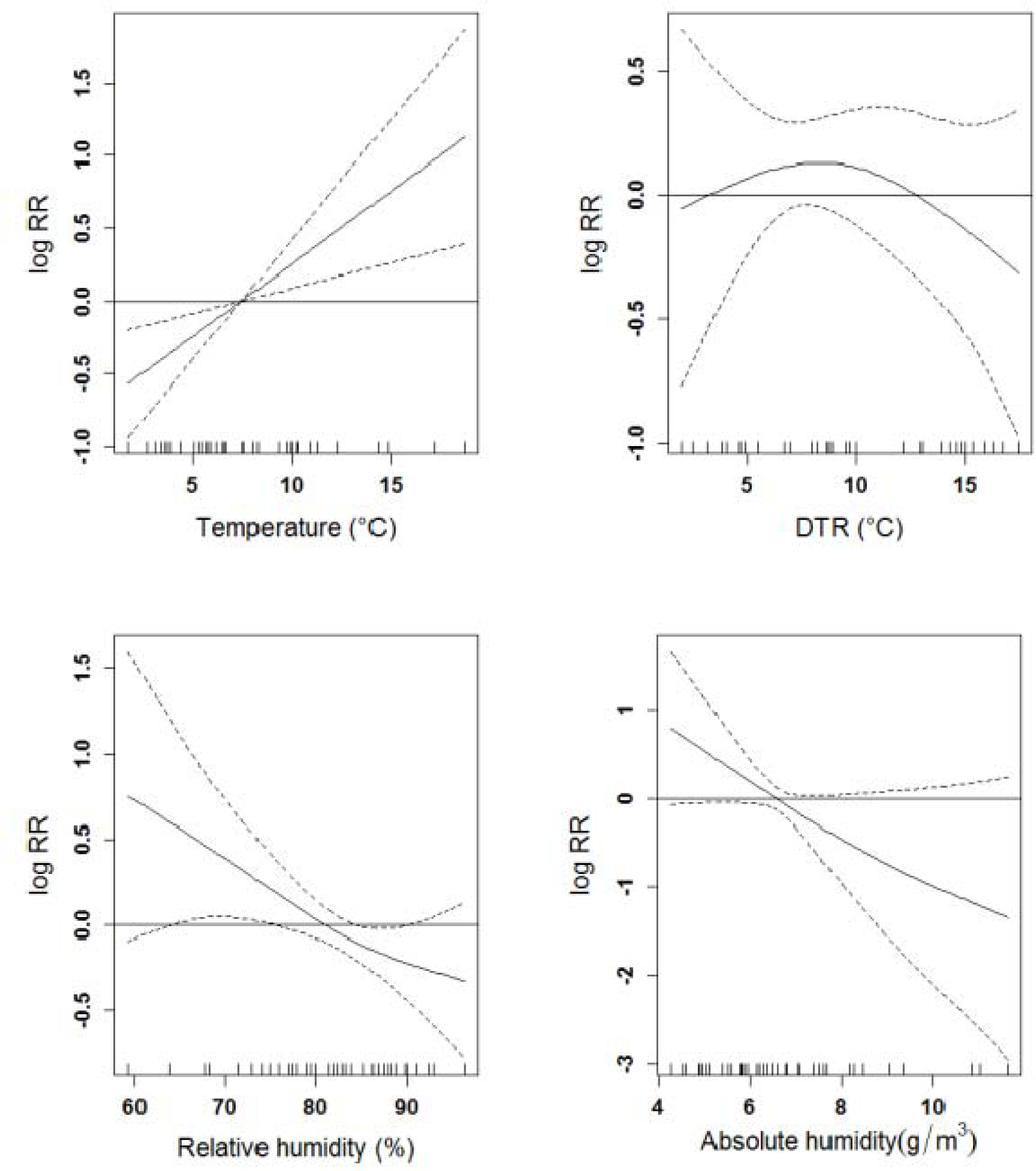
The exposure-response curves of meteorological factors and COVID-19 daily mortality counts in Wuhan, China, from 20 January to 29 February 2020. The X-axis is the concurrent day meteorological data, Y-axis is the predicted log relative risk (RR), is shown by the solid line, and the dotted lines represent the 95% confidence interval (CI). COVID-19, coronavirus disease; DTR, diurnal temperature range.

Fig. 3 displayed the percentage changes of COVID-19 mortality with per 1 unit increase in meteorological factor levels for different lag days in the models. DTR, temperature and absolute humidity were significantly associated with the increased/decreased COVID-19 mortality, after controlling the effects of air pollution and other factors. Each 1 unit increase in DTR was only associated with a 2.92% (95% CI: 0.61%, 5.28%) increase in COVID-19 mortality at lag 3. However, both per 1 unit increase of temperature and absolute humidity were related to the decreased COVID-19 mortality at lag 3 and lag 5, with the greatest decrease both at lag 3 [-7.50% (95% CI: -10.99%, -3.88%) and -11.41% (95% CI: -

**Fig. 3.**
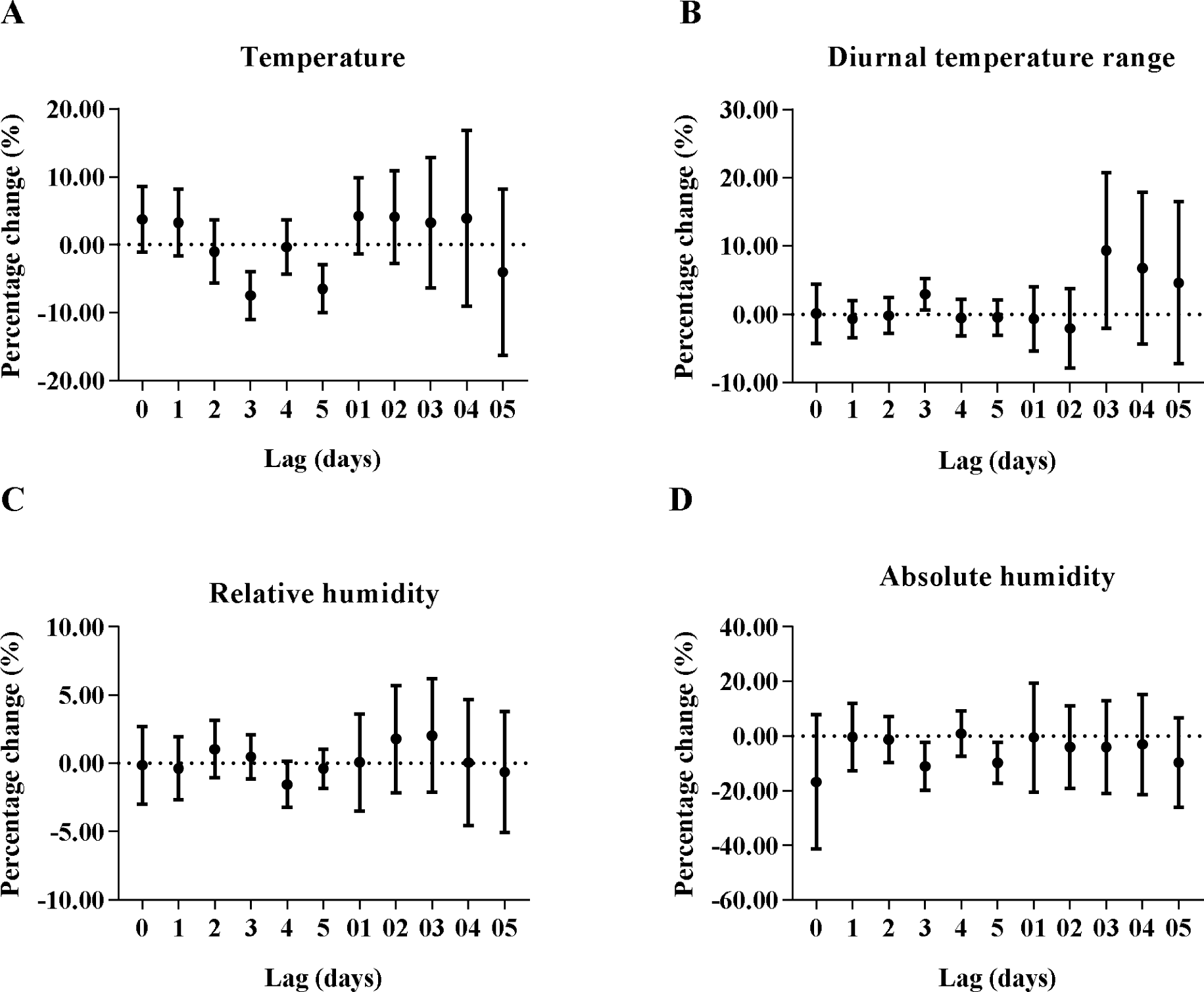
Percentage change (95% confidence interval) of COVID-19 daily mortality with per 1 unit increase in meteorological factors for different lag days in the models in Wuhan, China, from 20 January to 29 February 2020. COVID-19, coronavirus disease.

## 4. Discussion

The study was conducted in Wuhan during 20 January, 2020 to 29 February, 2020, in order to investigate the association between weather variables and daily death number of COVID-19. Significant positive effect of DTR on the daily mortality counts of COVID-19 was first identified through our study. It is also found that a significant negative association with COVID-19 mortality is observed for temperature and absolute humidity. The results indicate that the effects of DTR should also be paid attention on the mortality of COVID-19.

Our study demonstrated a negative association between COVID-19 case fatality and temperature, while a positive association for DTR. The results are consistent with the following investigations. Respiratory diseases mortality increased with decreasing temperature [24], and was strongly associated with low temperature [25-27]. While another study found that both cold and heat effects may have adverse impact on respiratory mortality [28]. Otherwise, the study conducted in 30 East Asian cities showed that increased DTR was associated with increased risk of mortality for respiratory and cardiovascular diseases [29]. In the cold season, the cumulative relative risk of non-accidental, respiratory and cardiovascular death increased at high DTR values in Tabriz [30]. A time-series study done in Shanghai on the effect of DTR and daily chronic obstructive pulmonary disease (COPD) mortality showing that each 1°C elevation in the 4-day moving average for DTR accounted for 1.25% of increased risk of COPD mortality [31]. A review for cold exposure and immune function reported that lower temperature may repress the immune function [32], particularly, our previous finding suggested that the phagocytic function of pulmonary alveolar macrophages declined under cold stress in vitro experiment [33]. Breathing cold air can lead to bronchial constriction, which may promote susceptibility to pulmonary infection [34]. Additionally, since SARS-CoV-2 is sensitive to heat, and high temperature makes it difficult to survive, not to mention the beneficial factors for virus transmission like indoor crowding and poor ventilation in cold days [35]. Also, cold temperature has been discovered to be associated with the reduction of lung function and increases in exacerbations for people with COPD [36]. In addition, DTR represents a stable measure of temperature, which is an indicator of temperature variability to evaluate effects on human health, including mortality and morbidity [37]. Besides, abrupt temperature changes may add the burden of cardiac and respiratory system causing cardiopulmonary events and high DTR levels may be a source of environmental stress [30]. Our findings highlight a need to strengthen the awareness of the effects of temperature and DTR on COVID-19 mortality.

Researchers confirmed that respiratory infection was enhanced during unusually cold and low humidity conditions [38], indicating low humidity may be also an important risk factor for respiratory diseases. Worse, a study with data of more than 25 years found that humidity was an important determinant of mortality and low-humidity levels may cause a large increase in mortality rates, potentially by influenza-related mechanisms [39], similar to a study carried out in the United States [40]. Consistent to these finding, our results also indicated that the risk of dying from COVID-19 decreased only with absolute humidity increasing. Breathing dry air could cause epithelial damage and/or reduction of mucociliary clearance, and then lead to render the host more susceptible to respiratory virus infection; The formation of droplet nuclei is essential to transmission, but exhaled respiratory droplets settle very rapidly at high humidity so that it is hard to contribute to influenza virus spread [41]. Moreover, the transmission of pandemic influenza virus is the most efficient under cold, dry conditions [42], and influenza virus survival rate increased markedly in accordance with decreasing of absolute humidity [43], which may be similar to coronavirus. Therefore, the increase of COVID-19 mortality may also be related to the lower humidity in winter.

However, many limitations should not be ignored. Firstly, there are some other factors may affect the COVID-19 mortality, such as population migration, government intervention and so on. Secondly, ecologic time-series study design was adopted in the study, which might exist ecologic fallacy. Furthermore, it is difficult to obtain the meteorological and air pollution data at the individual level. Nevertheless, this study showed that DTR might be associated with the death risk of COVID-19 in Wuhan, which deserves further investigation.

## 5. Conclusion

This is the first study to investigate the effects of temperature, DTR and humidity on daily mortality of COVID-19 in Chinese population by utilizing publicly accessible data on COVID-19 statistics and weather variables. Our finding shows that daily mortality of COVID-19 is positively association with DTR but negatively with relative humidity. In summary, this study suggests the temperature variation and humidity may be important factors affecting the COVID-19 mortality.

## Data Availability

Daily death number of COVID-19 were collected from the official website of Health Commission of Hubei Province, people's Republic of China (http://wjw.hubei.gov.cn/). The simultaneous daily meteorological and air pollutant data were obtained from Shanghai Meteorological Bureau and Data Center of Ministry of Ecology and Environment of the People’s Republic of China (http://www.datacenter.mee.gov.cn/), respectively.

## Abbreviations

COVID-19: coronavirus disease
Tem: temperature
DTR: diurnal temperature range
RH: relative humidity
AH: absolute humidity
PM_2.5_: particulate matter with aerodynamic diameter ≤2.5 μm
PM_10_: particulate matter with aerodynamic diameter ≤10 μm
NO_2_: nitrogen dioxide
SO_2_: sulfur dioxide
CO: carbon monoxide
O_3_: ozone
GAM: generalized additive model
CI: confidence interval
RR: relative risk

## Ethical Approval and Consent to participate

Not applicable.

## Consent for publication

Not applicable.

## Availability of supporting data

The datasets used and/or analyzed during the current study are available from the websites.

## Competing interests

The authors declare that they have no conflict of interest.

## Funding

This work was supported by the Novel Coronavirus Disease Science and Technology Major Project in Gansu Province.

## Author Contributions

Bin Luo, Jingping Niu, Yueling Ma and Yadong Zhao contributed to idea formulation, study design, data preparation, data analysis, reporting results, data interpretation, and writing of the manuscript. Jiangtao Liu and Jun Yan contributed to data preparation and data analysis. Xiaotao He, Bo Wang and Shihua Fu contributed to study design and interpretation of the data. All authors have seen and approved the final version.

## Acknowledgements

Not applicable.

## Authors’ information

^a^ Institute of Occupational Health and Environmental Health, School of Public Health, Lanzhou University, Lanzhou, Gansu 730000, People’s Republic of China. ^b^ Sexually Transmitted Disease and Acquired Immune Deficiency Syndrome Prevention Branch, Gansu Provincial Center for Disease Control and Prevention, Lanzhou, Gansu 730000, People’s Republic of China. ^c^ Department of General Surgery, the First Hospital of Lanzhou University, Lanzhou, Gansu 730000, People’s Republic of China. ^d^ Shanghai Typhoon Institute, China Meteorological Administration, Shanghai 200030, China. ^e^ Shanghai Key Laboratory of Meteorology and Health, Shanghai Meteorological Bureau, Shanghai, 200030, China

## Notes

### Competing Interest Statement

The authors have declared no competing interest.

